# Cellular and Humoral Immunity to Ebola Zaire Glycoprotein and Viral Vector Proteins Following Immunization with Recombinant Vesicular Stomatitis Virus-Based Ebola Vaccine (rVSVΔG-EBOV-GP)

**DOI:** 10.1101/2021.11.09.21266118

**Authors:** Vanessa Raabe, Lilin Lai, Juliet Morales, Yongxian Xu, Nadine Rouphael, Richard T. Davey, Mark J. Mulligan

## Abstract

While effective at preventing Zaire ebolavirus (EBOV) disease, cellular immunity to EBOV and vector-directed immunity elicited by the recombinant vesicular stomatitis virus expressing Ebola glycoprotein (rVSVΔG-EBOV-GP) vaccine remains poorly understood. Sera and peripheral blood mononuclear cells were collected from 32 participants enrolled in a prospective multicenter study [ClinicalTrials.gov NCT02788227] before vaccination and up to six months post-vaccination. IgM and IgG antibodies, IgG-producing memory B cells, and T cell reactivity to EBOV glycoprotein, vesicular stomatitis virus-Indiana strain (VSV-I) matrix protein, and VSV-I nucleoprotein were measured using ELISA, ELISpot, and intracellular cytokine staining, respectively. Eleven participants previously received a different investigational Ebola vaccine. All participants met positivity criteria for IgG antibodies to, and circulating IgG-producing memory B cells to, EBOV glycoprotein following rVSVΔG-EBOV-GP vaccination. Transient IgM and IgG antibody responses to VSV-I matrix protein (n=1/32 and n=0/32, respectively) and nucleoprotein (n=2/32 and n=1/32, respectively) were infrequently detected, as were IgG-producing memory B cells recognizing VSV-I matrix protein (n=3/31) and nucleoprotein (n=2/31). CD4+ and CD8+ T cell responses to EBOV glycoprotein were present in 15/32 and 19/32 participants at baseline and in 32/32 and 23/32 participants one month post-vaccination, respectively. CD4+ and CD8+ T cell responses to VSV-I matrix protein (n=17/32 and n=16/32, respectively) and VSV-I nucleoprotein (n=23/32 for both CD4+ and CD8+ responses) were common post-vaccination. T cell responses were predominantly mono-cytokine, except CD8+ responses to EBOV glycoprotein among heterologous Ebola vaccine-experienced participants and CD8+ responses to VSV-I nucleoprotein. Overall, rVSVΔG-EBOV-GP elicits robust humoral and memory B cell responses to EBOV glycoprotein in both Ebola vaccine-naïve and heterologous Ebola vaccine-experienced individuals and can generate vector-directed T cell immunity. Further research is needed to understand the significance of pre-existing vector-directed immunity on responses to booster doses of rVSVΔG-EBOV-GP and other rVSV-vectored vaccines.

## Introduction

Ebola virus disease is a continuing threat to human health since it was first identified in 1976, notably causing large-scale outbreaks in West Africa from 2014-2016 involving over 28,000 cases and 11,325 deaths and in the Democratic Republic of Congo from 2018-2020 with over 3400 cases and 2200 deaths [1, 2]. These large-scale epidemics accelerated the development and testing of Zaire ebolavirus (EBOV) vaccines, leading to European Medicines Agency and the U.S. Food and Drug Administration approval of a recombinant vesicular stomatitis virus expressing Ebola glycoprotein (rVSVΔG-EBOV-GP) vaccine in 2019 for the prevention of disease due to Zaire ebolavirus [3, 4]. This vaccine utilizes a replication-competent vesicular stomatitis virus – Indiana strain (VSV-I) from which the VSV-I glycoprotein (GP) gene has been removed and replaced by the EBOV GP gene. The efficacy of the rVSVΔG-EBOV-GP vaccine for prevention of disease due to EBOV was estimated from a ring-vaccination study during an Ebola outbreak to be 97.5% ten days or more post-vaccination and 88.1% at any time post-vaccination following administration of a single dose over a follow up period of approximately 11 months [5]. Ebola GP antibodies primarily mediate the protection conferred by the rVSVΔG-EBOV-GP vaccine although the vaccine can elicit both CD4+ and CD8+ T cell responses to EBOV GP [6, 7]. A combination of EBOV GP antibody titers ≥ 200 EU/mL plus a minimum 2-fold rise in EBOV GP antibody titers post-vaccination has been proposed as a vaccine-induced seroresponse in humans, although a minimum antibody titer necessary to confer protection has not been established [8]. CD4+ T cells play an important role in vaccine-induced antibody development, but not in protection at the time of EBOV challenge, while CD8+ T cells may play a minor role in protection [7, 9].

Knowledge gaps remain regarding cellular immunity to EBOV GP and vector-induced immunity elicited by rVSVΔG-EBOV-GP vaccination. Understanding vaccine vector-induced immunity is important to assess for any unintended, potentially adverse consequences associated with use of vectored vaccines in the setting of pre-existing vector immunity, such as modification of vaccine-induced target antigen immunity or the potential to enhance susceptibility to infection, as was observed among studies of adenovirus type 5 vectored HIV vaccine recipients [10-13], but also to understand the potential for vector-induced immunity to limit target antigen immune responses generated by subsequently administered vaccine doses utilizing the same vector. Human infection with VSV-I is rare and the prevalence of naturally acquired VSV-I antibodies is generally low with the exceptions of a few, geographically restricted regions [14]. The rVSVΔG-EBOV-GP vaccine can elicit anti-VSV-I antibodies in non-human primates, although these did not interfere with protection induced by a subsequently administered and similarly designed recombinant vesicular stomatitis virus (rVSV) vaccine expressing Lassa virus GP [15]. In retrospective analysis of a phase I clinical trial, transient, non-neutralizing antibodies directed against the VSV-I matrix (M) protein and T cell responses against the VSV-I nucleoprotein (NP) were elicited in a third of human rVSVΔG-EBOV-GP vaccine recipients [16].

Expanding our understanding of immunity elicited by the rVSVΔG-EBOV-GP vaccine against the target antigen and the VSV-I vector is critical for understanding the potential for long-term vaccine-induced protection against EBOV and the potential need for additional doses of rVSVΔG-EBOV-GP vaccine, or future doses of other rVSV-based vaccines, to successfully initiate or boost immunity to the target antigen. Although the rVSVΔG-EBOV-GP vaccine is currently approved as a single dose vaccine, the duration of protection following vaccination is unclear and clinical trials assessing the immunological benefit of booster doses of rVSVΔG-EBOV-GP vaccination are underway [ClinicalTrials.gov NCT02788227, NCT02876328]. Here we describe humoral, memory B cell, and T cell responses directed against Ebola GP, VSV-I M protein, and VSV-NP elicited in the first 6 months following rVSVΔG-EBOV-GP vaccine administration from a subset of volunteers participating in an open-label, phase 2 clinical trial to assess the durability of the immune response from rVSVΔG-EBOV-GP. Additionally, we described differences in immune responses to the rVSVΔG-EBOV-GP among individuals receiving an Ebola vaccine for the first time and those who previously received different Ebola vaccines.

## Materials and Methods

### Specimen Collection, Processing, and Storage

Additional blood specimens for this immunology substudy were obtained from a subset of participants enrolled in the PREPARE study [ClinicalTrials.gov NCT02788227]. Study participants received the rVSVΔG-EBOV-GP vaccine at their first study visit and were randomized at 18 months to a booster dose or no booster of the rVSVΔG-EBOV-GP vaccine. Written informed consent for the PREPARE study was obtained from all participants, which permitted the study site to collect additional research specimens at scheduled study visits. The PREPARE study was approved by the US Food and Drug Administration and the Emory University Institutional Review Board.

Additional sera, plasma, and peripheral blood mononuclear cells (PBMCs) were collected at baseline, prior to rVSVΔG-EBOV-GP vaccine administration, and one month, three months, and six months after initial rVSVΔG-EBOV-GP vaccination. Serum samples were obtained using serum-separating tubes containing silica clot activator and polymer gel (BD, 367983) and were processed by sitting at room temperature until clot formation was demonstrated followed by centrifugation. Plasma and PBMCs were obtained using CPT™ tubes with sodium heparin (BD #362753) following centrifugation. PBMCs were washed with phosphate-buffered saline (PBS), and cryopreserved in 90% fetal bovine serum (FBS) with 10% DMSO using a StrataCooler (Agilent) at -80°C, after which PBMCs were stored in liquid nitrogen prior to use. All samples were processed within two hours of collection. Aliquots of sera and plasma were stored at -80°C until ready for initial use and additional aliquots that underwent one freeze-thaw cycle were subsequently stored at -20°C.

### ELISA

Antibodies to Ebola GP, VSV-I GP, VSV-I NP, and VSV-I M proteins were assessed by indirect enzyme-linked immunosorbent assay (ELISA). Vesicular stomatitis virus GP is absent from the rVSVΔG-EBOV-GP vaccine and served as a control for natural VSV-I infection during the study period. In brief, Nunc Maxisorb plates (Fisher #439454) were coated with 1 µg/mL of recombinant Zaire ebolavirus glycoprotein minus trans-membrane region (IBT Bioservices 0501-015), VSV-I glycoprotein (Mudd-Summers strain, MyBioSource MBS1060254), VSV-I matrix protein (98COE North America strain, MyBioSource MBS1317470), or VSV-I nucleoprotein (Glasgow strain, MyBioSource MBS1026155) diluted with PBS and incubated at 4°C. Plates were blocked with PBS containing 0.05% Tween-20 (PBS-T), 5% dry milk, and 4% whey for 1 hour, after which diluted serum was added in three-fold dilutions starting at 1:100 and incubated at room temperature for 1 hours. Pooled serum from healthy volunteers participating an influenza vaccine study served as a negative control and serum from an Ebola convalescent patient (provided courtesy of the Centers for Disease Control and Prevention), murine anti-VSV glycoprotein antibodies (Sigma-Aldrich V5507) diluted 1:2000, murine anti-VSV matrix antibodies (Kerafast EB0011) diluted 1:2000, and murine anti-VSV nucleoprotein antibodies (Kerafast EB0009) diluted 1:2000 were used as positive controls. Plates were washed with PBS-T and incubated with horseradish peroxidase (HRP) goat anti-human IgG antibody (Jackson ImmunoResearch #109-036-098) diluted 1:10000, HRP goat anti-mouse IgG Fc (ThermoFisher A16084) diluted 1:20000, or HRP goat anti-human IgM antibody (Jackson ImmunoResearch #109-035-129) diluted 1:20000 for 1 hour. KPL SureBlue TM TMB Microwell Peroxidase Substrate (KPL 52-00-00) was added for 5 minutes, the reaction was stopped with 1N hydrochloric acid (VWR #BDH3202-2), and optical densities at 450 nm were read using a Biotek EL808 ELISA plate reader at room temperature. All samples were run in duplicate. We defined a positive antibody response to VSV-I proteins by an optical density (OD) at 450 nm at 1:100 dilution at least three standard deviations above the mean OD of all individuals at baseline and for Ebola glycoprotein, by an OD at 450 nm at 1:100 dilution at least three standard deviations above the mean OD of vaccine-naïve individuals at baseline.

### PBMC Thawing

Cryopreserved PBMCs were thawed in R10 containing DNase I (Roche #04716728001), washed, and resuspended in R10 with DNase I. Cell counting and viability assessment was performed using a Guava easyCyte counter (Luminex) per the manufacturer instructions. For intracellular cytokine staining, freshly thawed cells were rested overnight at an incubator at 37°C with 5% carbon dioxide and repeat counting and viability assessment were performed prior to use.

### IgG-producing Memory B Cell ELISpot

IgG-producing memory B cell (MBC) ELISpots were performed using recombinant Zaire ebolavirus glycoprotein minus trans-membrane region, VSV-I matrix protein, and VSV-I nucleoprotein on PBMCs at baseline and 6 months post-vaccination. In brief, cryopreserved PBMCs were thawed, stimulated with B-poly-S containing IL-2 and R848 (Cellular Technology Limited CTL-hBPOLYS-200) and 2-mercaptoethanol (Sigma-Aldrich M3148) in a 24 well flat-bottom cell culture plate (Fisher Scientific 07-200-740), and incubated for 5 days at 37°C. 96-well multiscreen-HA plates (Millipore MSHAN4B50) were coated with 2.5 µg/mL EBOV-GP, 2 µg/mL of VSV-I M protein, VSV-I NP, or donkey anti-human IgG (Jackson Immuno Research Laboratory 709-005-149) and stored at 4°C for 1-7 days prior to use. On day 6, plates were washed with PBS-T and blocked with RPMI 1640 with L-glutamine and HEPES (Fisher MT10041CV) with 10% fetal bovine serum (Hyclone SH30088.03) and penicillin and streptomycin solution (Cellgro 30-002-CI). Three-fold dilutions of PBMCs were added to the 96-well plates starting at 1:3 dilution for viral antigen wells and 1:30 dilution for anti-human IgG wells and incubated for 6 hours at 37°C. Plates were washed with PBS and PBS-T, and incubated overnight at 4°C with biotin-conjugated donkey anti-human IgG Fc antibody (Jackson Immuno Research Laboratory 709065098). After washing with PBS-T, samples were incubated with Avidin D conjugated with HRP (Vector Laboratories A-2004) for 1 hour at room temperature, plates were washed with PBS-T and PBS. 3-amino-9-ethylcarbazole substrate (Sigma A-5754) was added for 5 minutes and plates were washed with water and allowed to dry in the dark prior to being analyzed with an Immunospot S6 Universal Analyzer (Cellular Technology Limited). All samples were run in duplicate. To account for differences in the quantity of IgG-producing memory B cells present per million PBMCs, the number of antigen-specific spots per million PBMCs was divided by the total number of IgG-producing spots per million PBMCs and multiplied by 100 to produce an antigen-specific/total IgG-producing memory B cell percentage. An antigen-specific/total IgG-memory B cell percentage of ≥ 0.10% was considered a positive test.

### Intracellular Cytokine Staining

Intracellular cytokine staining of PBMCs collected at baseline and one month post-vaccination were assessed for production of interferon γ (IFN-γ), interleukin 2 (IL-2), and tumor necrosis factor α (TNF-α) from T cells stimulated with peptide pools spanning the Zaire ebolavirus GP (Mayinga strain, JPT PM-ZEBOV-GPMay), VSV-I M protein, or VSV-I NP. Peptides sequences to match the recombinant VSV-I M (MyBioSource MBS1317470) and NP (MyBioSource MBS1026155) antigens used for ELISA and ELISpot assays were custom prepared by Genscript as 15-mer peptides with 10-mer overlaps sequence and pooled to create a concentration of 200 µg/mL. Stimulated cells without antigen and staphylococcal enterotoxin B (Sigma-Aldrich S4881) served as negative and positive controls.

In brief, cryopreserved PBMCs were thawed and rested overnight as above. Cells were incubated with antigen-specific peptide pools, CD28 (BD #555725), and CD49d (BD #555501) for 6 hours at 37°C; brefeldin A and monensin (eBioscience #5537) were added after 2 hours of incubation. Cells were washed, stained with viability dye (Zombie Aqua, Biolegend #L423101), and fixated/permeabilized using Cytofix/Cytoperm (BD, #554722). Staining was performed using fluorescent-conjugated antibodies to CD3 (SP34-2, #562877), CD4 (L200, #560836), CD8 (RPA-T8, #555367), IL-2 (MQ1-17H12, #554567), and TNF-α (Mab11, #560679) from BD Bioscience and IFN-γ (4S.B3, #47731942) from eBioscience. Flow cytometry was performed on a LSRII (BD Bioscience) and data analysis was performed using FlowJo. A positive antigen-specific T cell response was defined as ≥ 0.01% of T cells producing at least one cytokine and ≥ 2-fold antigen-specific production of the same cytokine compared to the same visit negative control.

## Results

Blood samples were obtained from thirty-two participants enrolled in the PREPARE study. Samples were obtained at baseline prior to rVSVΔG-EBOV-GP vaccine administration, one month post-vaccination, and three months post-vaccination for all 32 participants and at six months post-vaccination for thirty-one participants. The median age of participants was 40 years (age range: 24 – 67 years) and 11/32 (34.3%) had previously received at least one heterologous investigational Ebola vaccine, either a replication-defective chimpanzee adenovirus 3 vector vaccine expressing Zaire Ebolavirus glycoprotein (ChAd3-EBO-Z) alone or ChAd3-EBO-Z followed by a modified vaccinia Ankara virus expression EBOV glycoprotein and other filovirus antigens (MVA-BN-Filo) [Table 1].

**Table 1:** Demographics of Study Participants

|  | Number of Participants (%)<br>N= 32 |
| --- | --- |
| Gender |  |
| Female | 17 (53.1%) |
| Male | 15 (46.9%) |
| Age (years) |  |
| 20-29 | 3 (9.4%) |
| 30-39 | 13 (40.6%) |
| 40-49 | 11 (34.4%) |
| 50-69 | 3 (9.4%) |
| ≥ 60 | 2 (6.3%) |
| Baseline Ebola Vaccine Status |  |
| Naïve | 21 (65.6%) |
| ChAd3-EBO-Z only | 2 (6.3%) |
| ChAd3-EBO-Z with MVA-BN-Filo booster | 9 (28.1%) |

ChAd3-EBO-Z: replication-defective chimpanzee adenovirus 3 vector vaccine expressing Zaire ebolavirus glycoprotein. MVA-BN-Filo: modified vaccinia Ankara virus expressing Zaire ebolavirus glycoprotein and other filovirus antigens (glycoproteins from Marburg virus, Sudan virus, and Tai Forest virus).

### Serology

Ebola GP IgG antibodies met positivity criteria (OD ≥ 0.341 at 1:100 dilution) in 7/11 (63.6%) of heterologous Ebola vaccine-experienced participants and no Ebola vaccine-naïve participants at baseline, in 31/32 (96.9%) participants one month post-vaccination, 32/32 (100%) participants three months post-vaccination, and 30/31 (96.8%) participants six months post-vaccination (Figure 1A). At baseline, the mean IgG OD to Ebola GP was significantly higher (p=0.03) in Ebola vaccine-experienced participants compared to Ebola vaccine-naï ve participants (mean OD 0.847 to 0.153; Figure 1B). Ebola GP IgG ODs rose following vaccination in all but one Ebola vaccine-experienced participant who had a very high baseline Ebola GP IgG OD of 3.05. Lower Ebola GP IgG OD values were observed among all participants ages 20-29 compared to older participants at all time points (baseline: p=0.026, 1 month: p=0.003, 3 months: p<0.001, 6 months: p<0.001) although no differences in OD values were observed when the analysis was restricted only to participants who were vaccine-naïve at study entry (baseline: p=0.89, 1 month: p=0.89, 3 months: p=0.95, 6 months: p=0.91).

**Figure 1:**
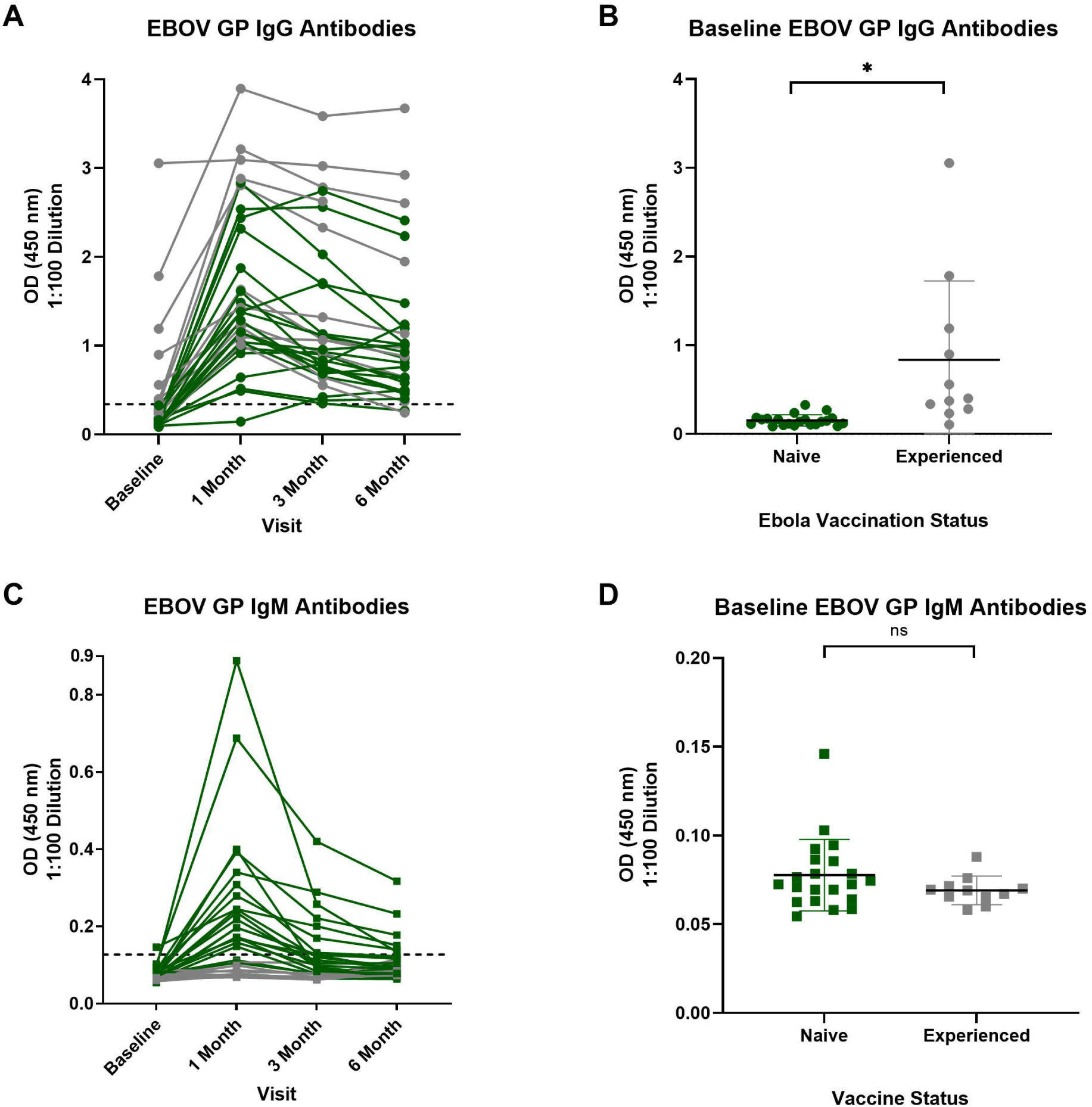
Antibody responses to Zaire ebolavirus glycoprotein by ELISA Baseline vaccine-experienced participants are shown in grey and vaccine-naïve participants are shown in green. A: EBOV GP IgG antibody optical densities at 1:100 dilution for each participant from baseline prior to rVSVΔG-EBOV-GP vaccination and one, three, and six months post-vaccination. The positive OD threshold is indicated by a dashed line. B: EBOV GP IgG antibody optical densities at 1:100 dilution prior to rVSVΔG-EBOV-GP vaccination for Ebola vaccine-naïve and heterologous Ebola vaccine-experienced participants, p=0.029. C: EBOV GP IgM antibody optical densities at 1:100 dilution for each participant from baseline and one, three, and six months post-vaccination. The positive OD threshold is indicated by a dashed line D: EBOV GP IgM antibody optical densities at 1:100 dilution at baseline for Ebola vaccine-naïve and heterologous Ebola vaccine-experienced participants, p=0.1. ns signifies p≥0.05, * signifies p<0.05.

IgM antibodies to EBOV GP were positive (OD ≥ 0.127 at 1:100 dilution) in 18/32 (56.3%) of participants one month post-rVSVΔG-EBOV-GP vaccination, 8/32 (25%) participants three months post-vaccination, and 6/31 (19.4%) at six months post-vaccination (Figure 1C). At baseline, no difference was observed between mean EBOV GP IgM ODs in Ebola vaccine-naïve) versus experienced participants (mean OD 0.078 vs 0.075; p=0.10, Figure 1D). Positive Ebola GP IgM ODs were observed only among Ebola vaccine-naïve participants and did not differ based on age or gender (data not shown). Among Ebola vaccine-naïve participants at enrollment, IgM responses were observed in 85.7%, 38.1%, and 28.6% at one month, three months, and six months post-vaccination, respectively.

No participants developed IgG antibodies to VSV-I M protein (OD ≥ 2.773 at 1:100 dilution; Figure 2A). One participant had IgM antibodies to VSV-I M protein (OD ≥ 0.173 at 1:100 dilution) detectable up to 3 months post-vaccination and one participant had an isolated, positive IgM VSV-I M protein level at baseline (Figure 2B). One participant developed a transient IgG responses to VSV-I NP (OD ≥ 2.359 at 1:100 dilution) at one month post-vaccination (Figure 2C). Two participants developed an IgM response to VSV-I NP (OD ≥ 0.297 at 1:100 dilution) one month post-vaccination while one participant had positive IgM antibodies to VSV-I NP detected only at baseline (Figure 2D). No participant developed IgG responses (OD ≥ 1.584) to VSV-I GP during the study period (Figure 2E).

**Figure 2:**
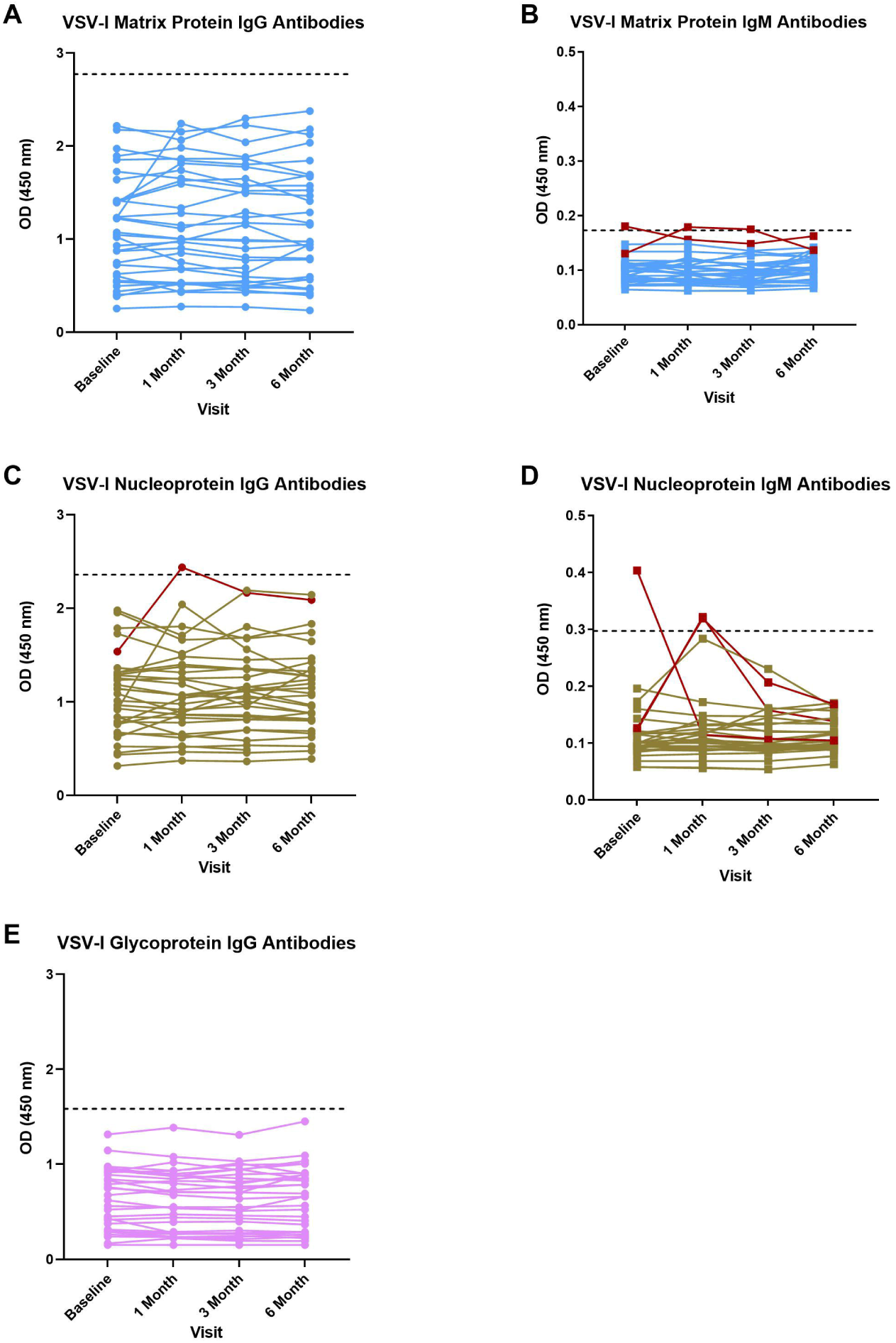
Antibody responses to vesicular stomatitis virus – Indiana strain matrix protein, nucleoprotein, and glycoprotein by ELISA Optical densities for each participant from baseline and one, three, and six months post-vaccination at 1:100 dilution for A) IgG antibodies to VSV-I matrix protein, B) IgM antibodies to VSV-I matrix protein, C) IgG antibodies to VSV-I nucleoprotein, D) IgM antibodies to VSV-I nucleoprotein, and E) IgG antibodies to VSV-I glycoprotein. The positive OD threshold is indicated by a dashed line and individuals with at least one value exceeding the positivity threshold are shown in red. The VSV-I matrix protein and nucleoprotein are present in the rVSVΔG-EBOV-GP vaccine while the VSV-I glycoprotein is absent from the vaccine.

### IgG-Producing MBCs

At baseline, 6 participants (19.4%), all of whom were Ebola-vaccine experienced, had detectable IgG-producing MBCs to EBOV GP while 31/31 (100%) had detectable EBOV GP antigen-specific IgG-producing MBCs at six months post-vaccination (Figure 3A; 3B). The percentage of EBOV GP specific MBCs was only weakly correlated with EBOV GP IgG ODs at 6 months post-vaccination (r=0.06). At baseline, 2 participants (6.5%) and 4 participants (12.9%) had detectable IgG-producing MBC responses to VSV-I NP and VSV-I M protein, respectively. Among participants positive for IgG-producing memory B cells to VSV-I antigens at baseline, VSV-I specific IgG-producing MBCs were no longer detectable at six months post-vaccination in all except one participant (3.2%) who had baseline detectable IgG-producing MBC responses to VSV-I M. Post-vaccination, two participants (6.5%) developed new IgG-producing MBC responses to VSV-I NP, both of whom also developed new IgG-producing MBC responses to VSV-I M protein (Figure 3C). One participant (3.2%) developed a new IgG-producing MBC response to VSV-I M protein post-vaccination, but not to VSV-I NP. None of the three participants with new IgG-producing MBC responses to VSV-I antigens post-vaccination had detectable serum antibodies to VSV-I M protein or NP following vaccination. IgG-producing MBC responses to VSV-I proteins were of a lower magnitude (0-0.20% antigen-specific/total IgG-producing MBCs) compared to the IgG-producing MBC response elicited by EBOV GP (0.11-2.68% antigen-specific/total IgG-producing MBCs).

**Figure 3:**
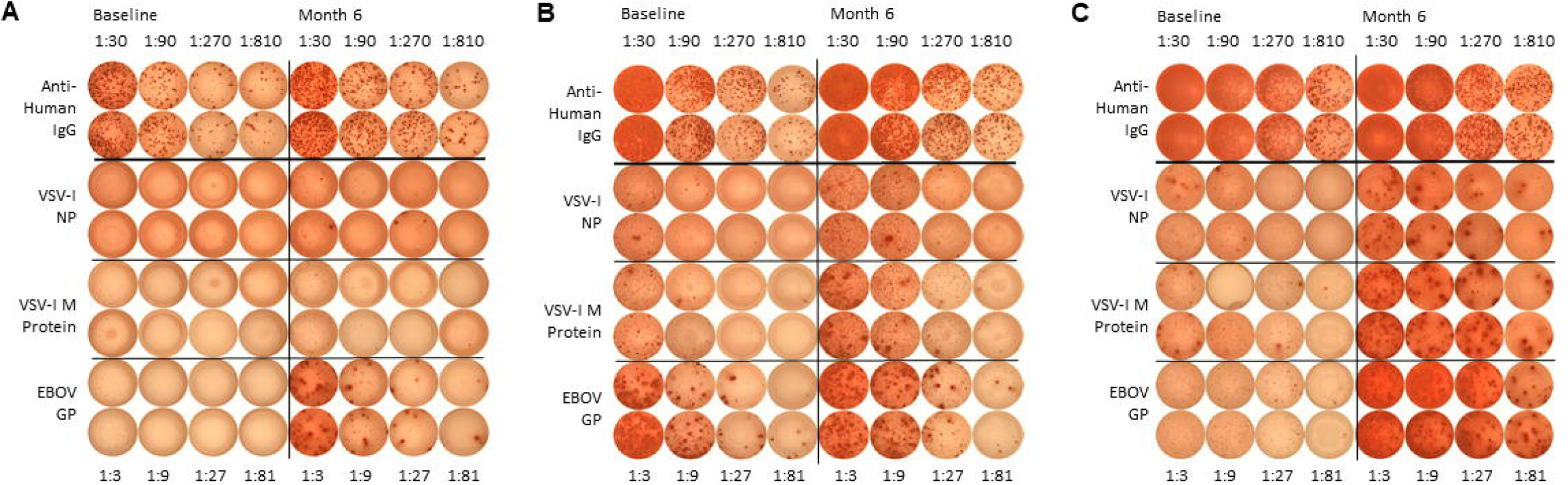
Representative IgG-producing memory B cell ELISpots at baseline and 6 months post-rVSVΔG-EBOV-GP vaccination A: A representative ELISpot from an Ebola vaccine-naïve participant demonstrating a new IgG-producing memory B cell (MBC) response at six months post-vaccination to EBOV GP only. B: A representative ELISpot from a heterologous Ebola vaccine-experienced participant with IgG-producing MBC responses to EBOV GP both at baseline and six months post-rVSVΔG-EBOV-GP vaccination. C: A representative ELISpot from an Ebola vaccine-naïve participant demonstrating new IgG-producing MBC responses at six months post-vaccination to EBOV GP, VSV-I M protein, and VSV-I NP.

### Intracellular Cytokine Staining

Positive CD4+ T cell responses following stimulation of PBMCs with peptides spanning the full-length EBOV GP were observed in 15 participants (46.9%, 9 Ebola vaccine-experienced, 6 Ebola vaccine-naïve) at baseline and 100% of participants one month post-vaccination. A significant increase in the mean percentage of CD4+ T cells producing IFN-γ post-vaccination was observed among Ebola vaccine-naïve participants (p=0.002) while Ebola vaccine-experienced participants experienced a significant increase (p=0.04) in the mean percentage of CD4+ T cells producing IL-2 (Figure 4A). Mean percentages of CD4+ T cells producing IL-2 and TNF-α upon stimulation with EBOV GP peptides were not different between Ebola vaccine-naïve and vaccine-experienced individuals at baseline or post-vaccination, while Ebola vaccine-experienced participants had higher mean percentages of CD4+ T cells producing IFN-γ than Ebola vaccine-naïve participants at baseline (0.020% to 0.004%, p=0.009) and but not one month post-vaccination (0.033% to 0.018%, p=0.05).

**Figure 4:**
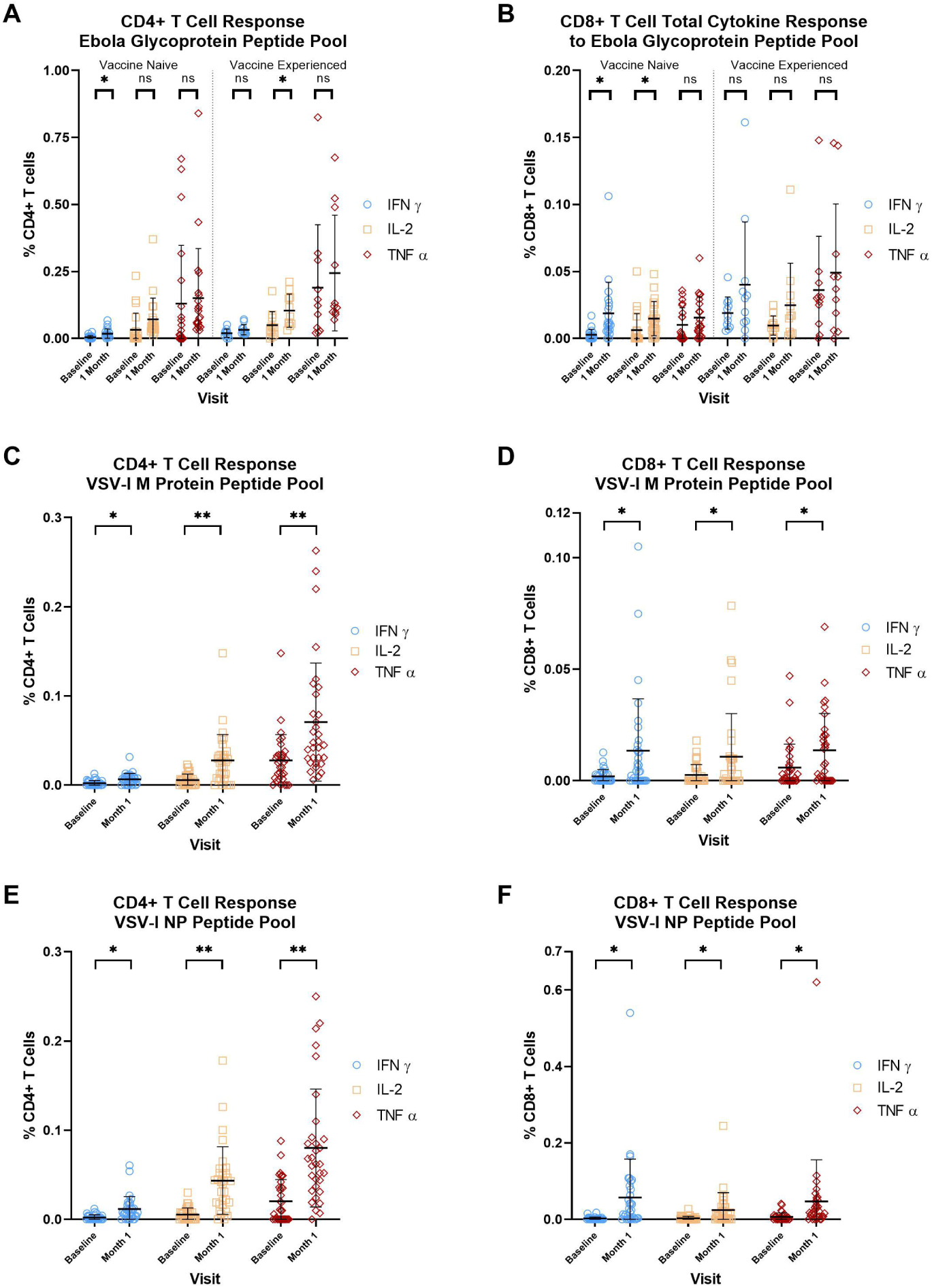
Intracellular cytokine staining for IFN-γ, IL-2, and TNF-α from CD4+ and CD8+ T cells Peripheral blood mononuclear cells from vaccinees at baseline and one month following rVSVΔG-EBOV-GP vaccination were assessed for production of IFN-γ, IL-2, and TNF-α from CD4+ or CD8+ T cells by intracellular cytokine staining using pooled peptides spanning the entire Zaire ebolavirus glycoprotein (A, B), VSV-I matrix protein (C, D), or VSV-I nucleoprotein (E,F). ns signifies p≥0.05, * signifies p<0.05, ** signifies p<0.001.

Nineteen participants (59.4%, 10 Ebola vaccine-experienced and 9 Ebola vaccine-naïve) had positive CD8+ T cell responses on stimulation with EBOV GP peptides at baseline while 23/32 participants (71.9%) had responses at one month post-vaccination. Significant increases in the mean percentage of CD8+ T cells producing IFN-γ (p=0.005) and IL-2 (p=0033) post-vaccination were observed among Ebola vaccine-naïve participants (Figure 4B). Ebola vaccine-experienced participants had a higher mean percentage of CD8+ T cells producing IFN-γ to EBOV GP peptides at baseline compared to vaccine-naïve participants (0.019% to 0.003%, p=0.001), otherwise no significant differences in the mean percentage of cytokine-producing CD8+ cells upon EBOV GP peptide stimulation were observed between Ebola vaccine-experienced and vaccine-naïve participants.

Stimulation with peptides spanning the VSV-I matrix protein led to significant increases in IFN-γ, IL-2, and TNF-α from both CD4+ T cells (IFN-γ: 0.22% to 0.67%; p=0.003; IL-2: 0.57% to 2.78%, p<0.001; TNF-α: 2.78% to 7.07%, p<0.001; Figure 4C) and CD8+ T cells (IFN-γ: 0.19% to 1.35%; p=0.009; IL-2: 0.26% to 1.08%, p=0.03; TNF-α: 0.59% to 1.37%, p=0.02; Figure 4D).Positive CD4+ and CD8+ T cell cytokine responses to VSV-I M protein were present in 2/32 (6.3%) and 6/32 (18.8%) of participants at baseline and 17/32 (53.1%) and 16/32 (50%) of participants at 6 months post-vaccination, respectively. Baseline T cell reactivity to VSV-I M protein was not associated with differences in the fold-change of Ebola GP IgG ODs at 1 month compared to baseline (Figure 5A, CD4+ T cells: p=0.71; CD8+ T cells: p=0.33).

**Figure 5:**
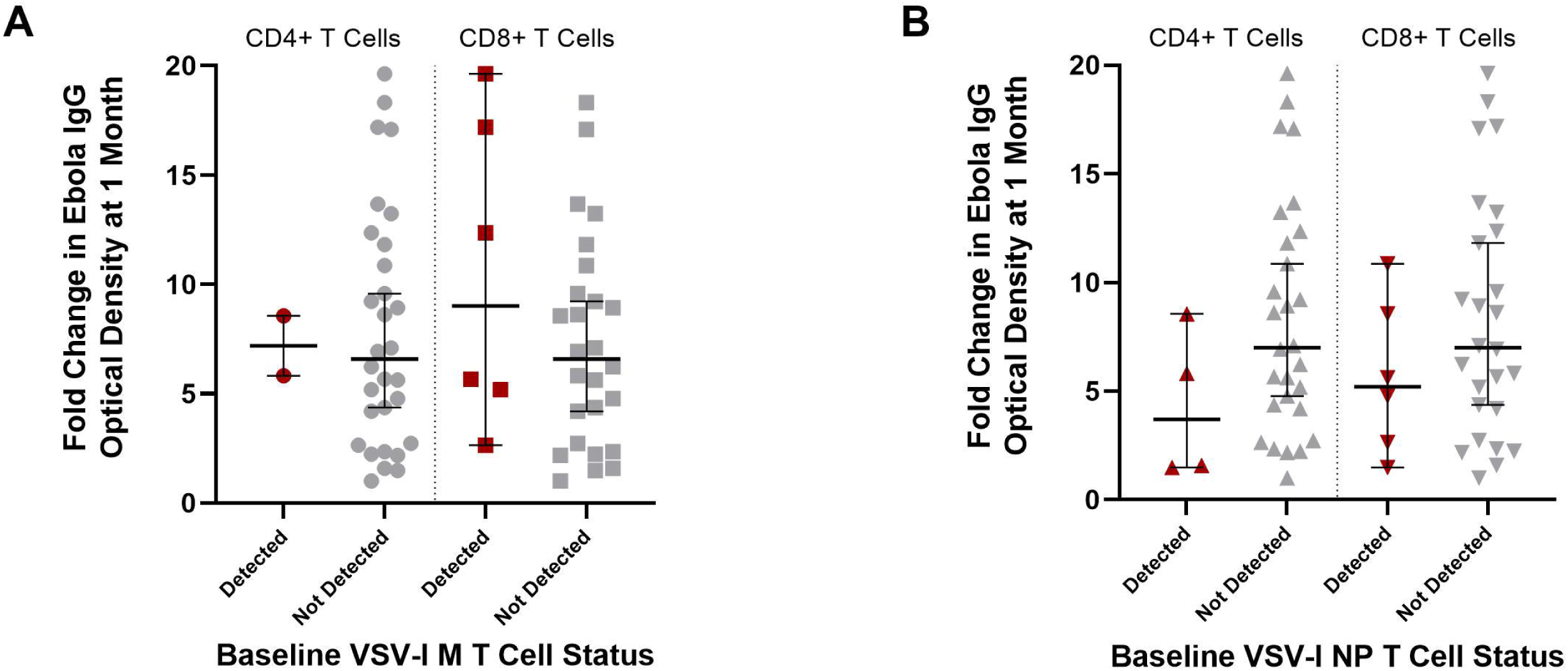
Fold change in Ebola IgG antibody optical densities at 1 month compared to baseline according the detectability of T cells recognizing VSV-I M (A) or NP (B) peptide pools at baseline. Median values with 95% intervals are shown.

Peptides from VSV-I NP similarly produced significant increases in IFN-γ, IL-2, and TNF-α from CD4+ T cells (IFN-γ: 0.21% to 1.16%; p=0.001; IL-2: 0.55% to 4.35%, p<0.001; TNF-α: 2.02% to 8.01%, p<0.001; Figure 4E) and CD8+ T cells (IFN-γ: 0.21% to 0.57%; p=0.004; IL-2: 0.28% to 2.42%, p=0.01; TNF-α: 0.66% to 4.69%, p=0.047; Figure 4F). Positive CD4+ and CD8+ T cell cytokine responses to VSV-I NP were present in 4/32 (12.5%) and 6/32 (18.8%) of participants at baseline and 23/32 (71.9%) and 21/32 (71.9%) of participants at 6 months post-vaccination, respectively. Neither baseline CD4+ or CD8+ reactivity to VSV-I NP were associated with differences in the fold-change of EBOV GP IgG ODs at 1 month compared to baseline (Figure 5B, CD4+ T cells: p=0.10; CD8+ T cells: p=0.16).

Post-vaccination CD4+ T cell responses to EBOV GP peptides were primarily mono-cytokine, while CD8+ T cell responses were predominantly mono-cytokine for Ebola vaccine-naïve participants and polyfunctional for heterologous Ebola vaccine-experienced participants (Figures 6A-D). Cellular responses to VSV-I Matrix protein were predominantly mono-cytokine for both CD4+ and CD8+ T cells (Figures 6E, 6F). CD4+ T cell responses to VSV-I NP were predominantly mono-cytokine while most CD8+ T cellular responses were polyfunctional (Figures 6G, 6H).

**Figure 6:**
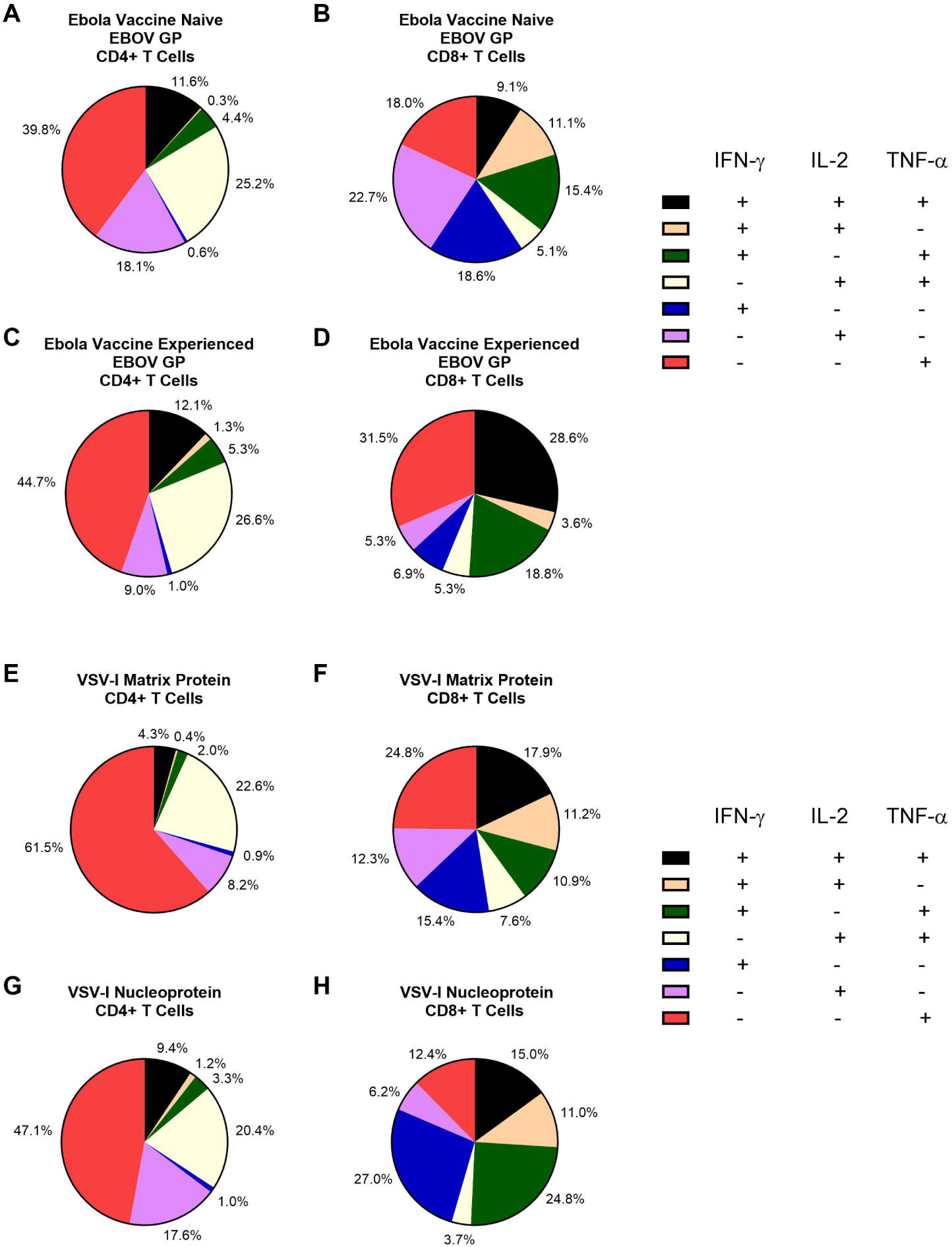
Pie charts representing the functionality of CD4+ or CD8+ T cells at one month post-rVSVΔG-EBOV-GP vaccination in response to peptides from Zaire ebolavirus glycoprotein, VSV-I matrix protein, or VSV-I nucleoprotein. Pie charts representing the mean functionality of CD4+ T cells (A, C, E, G) and CD8+ T cells (B, D, F, H) at one month following rVSVΔG-EBOV-GP vaccination to EBOV GP for Ebola vaccine-naïve (A, B, n=21) or Ebola vaccine-experienced (C, D, n=11) participants, and to VSV-I matrix protein (E,F, n=32) and VSV-I NP (G, H, n=32) among all participants. Due to rounding, the total mean percentage of cells of each functionality may not add up precisely to 100%.

## Discussion

Among the 32 participants in this study, all met our definition for a positive Ebola GP IgG response by three months post-rVSVΔG-EBOV-GP vaccination, including 4 heterologous Ebola vaccine-experienced individuals who did not meet positivity criteria at baseline. To our knowledge, this is the first study to describe the use of the rVSVΔG-EBOV-GP vaccine among prior heterologous Ebola vaccine recipients. Notably, the rVSVΔG-EBOV-GP vaccine induced increased EBOV GP IgG ODs in the majority of vaccine-experienced recipients, raising the potential for the rVSVΔG-EBOV-GP vaccine to be used as a booster for recipients of heterologous Ebola vaccines with waning immunity. Failure to increase Ebola IgG OD on ELISA following rVSVΔG-EBOV-GP vaccine administration only occurred in a single heterologous Ebola vaccine-experienced individual who exhibited high baseline ODs for EBOV GP IgG antibodies well above the study-defined positivity threshold. Among participants in a previous clinical trial that administered two doses of rVSVΔG-EBOV-GP vaccine twenty-eight days apart, only 12% of participants had detectable plasma rVSVΔG-EBOV-GP virus three days after the second dose of vaccination, compared to 95% after the first dose, suggesting vaccine-induced immunity inhibits the ability of the vaccine virus to replicate with subsequent doses [17]. We hypothesize the high baseline level of pre-existing EBOV GP IgG antibodies in the participant whose Ebola GP IgG OD did not increase post-rVSVΔG-EBOV-GP vaccination were responsible for neutralizing the vaccine virus prior to the establishment of successful rVSVΔG-EBOV-GP virus infection in this individual, however since the PREPARE protocol did not include early post-vaccination time points, we cannot definitively assess whether this is the case. The failure of this individual to respond to the rVSVΔG-EBOV-GP vaccine suggests further research is needed to understand the role of pre-existing EBOV GP IgG antibodies and their potential interference with the rVSVΔG-EBOV-GP vaccine to understand which populations are most likely to benefit from booster doses of rVSVΔG-EBOV-GP.

IgM antibodies to EBOV GP generated by the rVSVΔG-EBOV-GP vaccine have been demonstrated to provide up to 50% of antibody-mediated neutralization of EBOV, and therefore may contribute to early vaccine-mediated protection from disease [18]. IgM antibodies to EBOV GP were detectable only among participants who were Ebola vaccine-naïve prior to administration of the rVSVΔG-EBOV-GP vaccine. In contrast, IgM was the primary antibody type identified four weeks following two doses of rVSVΔG-EBOV-GP vaccine given twenty-eight days apart [18]. The differences in the observed results could be due to longer durations between Ebola vaccine doses among Ebola vaccine-experienced participants in this study or the type of Ebola vaccine used as the priming dose. Alternatively, it is possible that heterologous Ebola vaccine-experienced participants in this study mounted transient IgM antibody responses to rVSVΔG-EBOV-GP vaccine that resolved prior to the first post-vaccination visit one month later. Among Ebola naïve participants at baseline, IgM positivity declined after the one month visit, but was still detectable in nearly one-fifth of participants at six months post-vaccination. Prolonged EBOV GP IgM positivity following rVSVΔG-EBOV-GP vaccination have been noted in other studies, therefore caution should be used when interpreting diagnostic tests for EBOV that include EBOV GP IgM antibodies among rVSVΔG-EBOV-GP vaccinees [18, 19].

While the duration of protection against EBOV conferred by the rVSVΔG-EBOV-GP vaccine remains unclear, we demonstrated that the VSVΔG-EBOV-GP vaccine generates B cell memory to EBOV GP. All participants had detectable, circulating IgG-producing memory B cells recognizing EBOV GP following vaccination. Whether this immunological memory will be sufficient to provide complete protection against EBOV disease in the setting of waning serum antibody levels over time has yet to be determined.

Vaccine vector-directed humoral immune responses to the VSV-I matrix protein and nucleoprotein were uncommon among vaccine recipients. No participants developed IgG antibodies directed at the VSV-I matrix protein, in contrast to 28% of rVSVΔG-EBOV-GP vaccinees with detectable, although transient, IgG antibodies by Poetsch et al [16], and only one participant developed transient IgG antibodies to the VSV-I nucleoprotein. Likewise, detectable IgG-producing memory B cells to VSV-I proteins were low. Taken together, our data indicate that most rVSVΔG-EBOV-GP vaccinees do not acquire humoral immunity to viral vector proteins after one dose of rVSVΔG-EBOV-GP vaccine. Antibodies to VSV-I matrix protein induced by the rVSVΔG-EBOV-GP vaccine have been shown to be non-neutralizing *in vitro*, but the neutralizing capacity of VSV-I nucleoprotein antibodies in humans has not been assessed to our knowledge [16]. In a non-human primate model, antibodies to whole VSV were induced by a rVSV vaccine expressing EBOV GP, but did not impair protection from a subsequently given rVSV vaccine expressing Lassa virus GP, although only three primates were challenged [15]. Whether or not anti-VSV antibodies negatively impair responses to booster doses of rVSVΔG-EBOV-GP vaccine, or future doses other replication-competent, rVSV-based vaccines, in humans remains to be determined.

Similar to previous studies, we found the rVSVΔG-EBOV-GP vaccine can elicit CD4+ and CD8+ T cells to EBOV GP producing IFN-γ and IL-2 [6]. Production of IFN-γ and IL-2 T cell responses to EBOV GP may be associated with the development of symptoms such as arthralgias, fatigue, headache, and myalgias following rVSVΔG-EBOV-GP vaccination [20]. In this study, the cytokine profile produced by T cells differed between Ebola vaccine-experienced and vaccine-naïve individuals. EBOV GP CD8+ T cell responses to EBOV GP were more polyfunctional in nature in this study compared to those documented by Dahlke et al., especially among heterologous Ebola vaccine-experienced participants [6]. Interestingly, some of the Ebola vaccine-naïve individuals met our criteria for detectable T cell responses to EBOV GP peptides prior to vaccination, which may represents non-specific epitope cross-reactivity in the setting of seronegativity to EBOV GP. Cytokine production from CD8+ T cells stimulated with EBOV GP peptide pools prior to rVSVΔG-EBOV-GP vaccination was also observed in some participants studied by Dahlke et al. [6]. Although we cannot rule out immunological interference between pre-existing T cells recognizing EBOV GP with vaccine-induced EBOV GP antibody responses, we suspect these do not cause clinically significant interference with the rVSVΔG-EBOV-GP vaccine-induced target antigen humoral response given 100% of participants in this study met our criteria for a positive EBOV GP IgG response by 3 months post-vaccination despite 46.9% and 59.4% of participants having CD4+ and CD8+ T cells, respectively, capable of producing cytokines in response to EBOV GP peptide pools at baseline.

T cell responses to peptides from VSV-I proteins were uncommon at baseline among participants, but were relatively common at one month post-vaccination, particularly to the VSV-I nucleoprotein. The VSV-I nucleoprotein is the most common target for CD8+ T cell responses in mice infected with VSV-I [21], although no data exists on cellular immunity in humans infected with VSV-I to our knowledge. The proportion of participants with CD8+ T cell responses to VSV-I NP peptides in this study, 71.9%, is higher than that reported in a retrospective analysis of a phase I rVSVΔG-EBOV-GP clinical trial using three different vaccine doses, in which 36% of vaccinees demonstrated CD8+ T cell responses to VSV-I nucleoprotein [16]. Interestingly, we observed more polyfunctional VSV-I nucleoprotein-specific CD4+ and CD8+ T cells in our study than had previously been reported following vaccination, where predominantly mono-cytokine production of TNF-α was observed [16]. These vector-directed T cell responses potentially could negatively affect the immune response to booster doses to rVSV-based vaccines. We did not see differences in the fold-change of the Ebola GP IgG optical density at 1 month post-vaccination between the participants in this study with and without T cells responses to VSV-I proteins at baseline, however, this should be interpreted cautiously in the setting of the small number of participants with baseline T cell recognition of VSV-I proteins. The main PREPARE trial itself, from which our immunology substudy is derived, provides an opportunity to assess the impact of the vector-directed immune responses elicited by the rVSVΔG-EBOV-GP vaccine on the response to future doses, as participants are randomized to receive or not receive a booster dose of rVSVΔG-EBOV-GP vaccine eighteen months after the original dose.

Limitations of the study include the small number of participants assessed, especially of heterologous Ebola vaccine-experienced individuals. The Ebola vaccine-experienced individuals had previously received vaccine constructs other than the rVSVΔG-EBOV-GP vaccine, therefore, the immune responses among Ebola vaccine-experienced individuals reported in this study may not be reflective of vaccine responses provided by homologous prime and boost with the rVSVΔG-EBOV-GP vaccine. No early time points prior to one month post-vaccination were scheduled in the parent study to assess whether pre-existing EBOV GP IgG antibodies among Ebola vaccine-experienced participants altered the course of rVSVΔG-EBOV-GP vaccine viremia or the development of a circulating follicular helper T cell response post-vaccination, which correlates with EBOV GP antibody titers 4 weeks after vaccination [22]; important areas for future research to determine who may be most likely to benefit from rVSVΔG-EBOV-GP vaccine booster doses. Given the lack of data on immunity to VSV-I infection in humans and the absence of meaningful cut-offs for antibody titers, we presented data in optical density format to be comparable with previously reported studies and selected our lowest ELISA dilution as the most comprehensive for analysis, although we suspect the wide distribution of IgG ODs to all VSV-I proteins at baseline by ELISA indicates non-specific antigen cross-reactivity with non-VSV-I antibodies, which has previously been reported [23].

In conclusion, the rVSVΔG-EBOV-GP vaccine frequently elicits EBOV-GP-specific IgG antibody and IgG-producing memory B cell responses, and CD4+ and CD8+ T cell responses to EBOV GP in both Ebola vaccine-naïve and experienced individuals. T cell responses to VSV-I proteins are commonly elicited by rVSVΔG-EBOV-GP, although vaccine vector-directed humoral and IgG-producing memory B cell responses are rare. Additional research is needed to understand the implications of vaccine-elicited immunity, both to the vector and to target antigens, on the immune response to booster doses of the rVSVΔG-EBOV-GP vaccine.

## Data Availability

All data produced in the present study are available upon reasonable request to the authors.

## Abbreviations

ChAd3-EBO-Z: replication-defective chimpanzee adenovirus 3 vector vaccine expressing Zaire ebolavirus glycoprotein
EBOV: Ebola virus
GP: glycoprotein
M: Matrix protein
MBC: Memory B cell
MVA-BN-Filo: modified vaccinia Ankara virus expression Zaire ebolavirus glycoprotein and other filovirus antigens
NP: Nucleoprotein
PREPARE: The Multicenter Study of the Immunogenicity of Recombinant Vesicular Stomatitis Vaccine for Ebola-Zair for Pre-Exposure Prophylaxis in Individuals at Potential Occupational Risk for Ebola Virus Exposure
rVSV: Recombinant vesicular stomatitis virus
rVSVΔG-EBOV-GP: Recombinant vesicular stomatitis virus Ebola vaccine
VSV-I: Vesicular stomatitis virus Indiana strain

## Acknowledgements

We also wish to acknowledge the main study team at the NIH Clinical Center and at the University of Minnesota Data Center under whose auspices the PREPARE trial has been conducted. We would also like to thank our study participants for their volunteerism and acknowledge the important contributions of the following Emory Hope Clinic staff members: Dawn Battle, Mary Bower, Justin Colwell, Rijalda Deovic, Tamera Franks, Tigisty Girmay, Mari Hart, Christopher Huerta, Hannah Huston, Sara Jo Johnson, Brandi Johnson, Dean Kleinhenz, Pam Lankford-Turner, Hollie Macenczak, Regina Mosley, Eileen Osinski, Michele McCullough, Divyanshu Raheja, Brittany Robinson, Sirajud-Deen Talib and Juton Winston.

